# Racial and ethnic associations with interstitial lung disease and healthcare utilization in patients with systemic sclerosis

**DOI:** 10.1101/2024.02.02.24302197

**Authors:** Ann-Marcia C. Tukpah, Jonathan A. Rose, Diane L. Seger, Gary Matt Hunninghake, David W. Bates

**Affiliations:** Division of Pulmonary and Critical Care Medicine, Department of Medicine, Brigham and Women’s Hospital, Boston, MA; Division of General Internal Medicine and Primary Care, Department of Medicine, Brigham and Women’s Hospital, Boston, MA

**Keywords:** scleroderma, pulmonary fibrosis, emergency department, hospitalization, race/ethnicity, electronic health record

## Abstract

**Rationale:** Racial and ethnic differences in presentation and outcomes have been reported in systemic sclerosis (SSc) and SSc-interstitial lung disease (ILD). However, diverse cohorts and additional modeling can improve understanding of risk features and outcomes, which is important for reducing associated disparities.

**Objective(s):** To determine if there are racial/ethnic differences associated with SSc-ILD risk and age; time intervals between SSc and ILD, and with emergency department (ED) visit or hospitalization rates.

**Methods:** A retrospective cohort study using electronic health record data from an integrated health system, over a 5.5 year period was conducted using clinical and sociodemographic variables, models were generated with sequential adjustments for these variables. Logistic regression models were used to examine the association of covariates with ILD and age at SSc-ILD. Healthcare outcomes were analyzed with complementary log-log regression models.

**Results:** The cohort included 756 adults (83.6% female, 80.3% non-Hispanic White) with SSc with a mean age of 59 years. Overall, 33.7% of patients in the cohort had an ILD code, with increased odds for Asian (odds ratio [OR], 2.59; 95% confidence interval [CI], 1.29, 5.18; *P*=.007) compared to White patients. The age in years of patients with SSc-ILD was younger for Hispanic (mean difference, −6.5; 95% CI, −13, −0.21; *P* = 0.04) and Black/African American patients (−10; 95% CI −16, −4.9; *P* <0.001) compared to White patients. Black/African American patients were more likely to have an ILD code before an SSc code (59% compared to 20.6% of White patients), and had the shortest interval from SSc to ILD (3 months). Black/African American (HR, 2.59; 95% CI 1.47, 4.49; *P*=0.001) and Hispanic patients (HR 2.29; 95% CI 1.37, 3.82; *P*=0.002) had higher rates of an ED visit.

**Conclusion:** In this study, SSc-ILD presentation and outcomes differed by racial/ethnic group (increased odds of SSc-ILD, younger age at SSc-ILD, and preceding diagnosis with respect to SSc, rates of ED visit), some of which was attenuated with adjustment for clinical and sociodemographic characteristics. Differing presentation may be driven by social drivers of health (SDOH), autoantibody profiles, or other key unmeasured factors contributing to susceptibility and severity.

---

Systemic sclerosis (SSc) is a chronic autoimmune disease characterized by immune dysregulation, vasculopathy, and multiorgan fibrosis, with lung disease (interstitial lung disease or ILD) and/or pulmonary arterial hypertension (PAH)) as the leading cause of death.^1,2^ SSc-ILD is heterogeneous in progression and outcomes,^3^ significant pulmonary function decline^4^ and ILD development occur early.^5^ Therefore, timely identification of SSc-ILD is essential.^6,7^

Numerous studies suggest that race and ethnicity may be important factors in SSc-ILD presentation and outcomes. Racial/ethnic differences in age at SSc diagnosis have been reported in observational SSc cohorts,^8–10^ and age at SSc-ILD in clinical trial participants.^11^ Furthermore, the differential prevalence of SSc-ILD is demonstrated within multi-racial/ethnic cohorts.^12–14^ Black/African American race has been associated with a lower forced vital capacity (FVC),^15^ an increased odds of restrictive lung disease (RLD),^16^ and pulmonary fibrosis (PF).^9^ Despite these findings, some of these results are limited by sample size and SSc subtype,^15^ or lung disease severity,^9^ adjustment for important covariates,^14^ and the numbers of different racial/ethnic groups compared.^12^ SSc-ILD is known to be a principal cause of hospitalization,^6^ with increased healthcare utilization in patients with ILD,^17^ but there has been limited assessment of the impact of racial/ethnic differences on clinical outcomes such as emergency department (ED) visits,^18^ and hospitalizations.^19–21^ Further inquiry and analyses to expand understanding of possible informative traits of patient groups, and their impact on variability of presentation and trajectory can improve overall outcomes.

We hypothesized that among patients with SSc there would be important differences among racial/ethnic groups in ILD incidence, age at presentation, timing with respect to a SSc diagnosis, and important outcomes such as ED visits and hospitalizations. To test these hypotheses, we assessed an electronic health record (EHR) based cohort of patients with SSc from Mass General Brigham (MGB), an integrated multi-hospital healthcare system.^22^

## Methods

### Study Population and Variables

We performed a retrospective cohort study and selected study participants from MGB integrated health-system databases: the Research Patient Data Registry (RPDR) and Enterprise Data Warehouse (EDW). The cohort of 788 patients with SSc was created using a previously published algorithm in which patients with >=2 SSc International Classification of Diseases, Tenth Revision (ICD-10) codes were identified from January 1, 2016 through June 30, 2021.^22^ We identified patients with ILD using >=1 ILD ICD-10 codes (Table E1), sourced from encounters, billing, and the problem list. The first instance that the code was documented during the study period was considered the index date for SSc, and first ever code for ILD (specifically the earliest date as it could occur prior to SSc). Among the 788 patients, 756 who had an ILD code (or did not have a code) before or during the study period were included in the analyses (exclusions: n=31 with ILD code after the study period and n=1 American Indian or Alaska Native patient). Hereafter, SSc and ILD ICD-10 codes are referred to as SSc or ILD diagnosis respectively. Age at SSc and SSc-ILD were determined by age at index code use (if age at SSc-ILD preceded SSc age, it is denoted as negative).

Data elements including demographics (self-reported race/ethnicity), Charlson Comorbidity Index (CCI), medications, laboratory data, ED and hospitalization visits, median income imputed by zipcode in 2018, education, insurance, and employment status were collected. Race/ethnicity (recorded as “Black or African American” and, reported as Black/African American; Hispanic ethnicity is inclusive of all racial groups) data were missing for 34 patients, further chart review provided information for an additional 10 patients. Organ system involvement was based on selected ICD-10 codes for gastro-esophageal reflux disease (GERD), esophageal disease, Raynaud, pulmonary hypertension (PHTN), chronic kidney disease (CKD) and acute kidney injury (AKI). Ever order of a medication was determined by any prescription order available in our system during the study period but does not represent dispense nor use history.

Missing rate for variables were race/ethnicity (n=21, 2.8%), 2018 median family income (n=17, 2.2%), smoking (n = 290, 38.3%), education (n=130, 17.2%), employment (n=44, 5.8%). Missing data were imputed using Multiple Imputation by Chain Equations^23^ (MICE, version 3.15.0) with the predictive mean matching method for continuous median income, logreg method for binary employment and education, and polytomous regression method for multi-level categorical variables (race/ethnicity, smoking^24^). Five iterations with 25 imputed datasets were created and estimates were pooled for the regression models for each outcome (ILD, age at SSc/SSc-ILD, ED visit, Hospitalization) using Rubin’s rules.

### Statistical Analyses

We analyzed differences in patient characteristics between patients with ILD and those without using student T-test for continuous variables (after normality diagnostics, q-q plot visualization), Wilcoxon rank-sum test for continuous variables not normally distributed, and Fisher’s exact tests for categorical variables. Time distribution comparisons across groups in Ridgeline plots were conducted with the Kruskal-Wallis test. Multivariable linear or logistic regression assessed the association between age at SSc/SSc-ILD, and ILD outcome, using selected social drivers of health (SDOH: income, education, insurance, employment) and clinical covariates. Each ED visit and hospitalization were collapsed into the binary outcome of having at least 1 ED visit or hospitalization during the study period and used in the univariate and multivariable complimentary log-log binary regression models with the outcome as a rate. To account for varying person-time and follow-up, an offset of the log of patient-days was included. Sensitivity analyses were conducted 1) to assess associations in the restricted condition of using the ILD ICD-10 code sourced from the patient’s problem list (instead of any source), which could represent more clinician certainty and 2) to compare hospitalization rates for a pre-COVID-19 period (January 2016-February 2020), during-COVID-19 period (March 2020-June 2021), and overall study period. P-values, 2-sided, less than <0.05 were considered statistically significant, and 95% confidence interval were presented for proportions and regression analyses. Analyses were performed with R software (The R Foundation for Statistical Computing, version 4.2.2) using RStudio (version 2022.12.0+353). The study was approved by MGB Institutional Review Board 2021P001697.

## Results

### Demographic, Socioeconomic and Clinical Variables

Of the 756 SSc patients, 632 were women (83.6%) and 124 men (16.4%). The cohort was predominantly White (n=607, 80.3%) with a mean age of 59.4 years old at SSc index date. The sociodemographic and clinical characteristics of the cohort, comparing those with and without ILD are shown in Table 1.

**Table 1.** Comparison of baseline characteristics of patients with systemic sclerosis, with and without interstitial lung disease ICD-10 code (before or during study period)

| **Demographics** | **SSc-ILD**<br>**(n= 255)** | **SSc without ILD**<br>**(n= 501)** |
| --- | --- | --- |
| Age at index SSc code, years, mean ± SD | 58.4±14.1 | 59.9±14.9 |
| Female | 205 (80.4) | 427(85.2) |
| Race/ethnicity | 194 (76) | 413 (82.4) |
| White | 22 (8.6) | 23 (4.6) |
| Black/African American | 16 (6.3) | 30 (6) |
| Hispanic | 20 (7.8) | 17 (3.4) |
| Asian | 3 (1.1) | 18 (3.6) |
| Unknown |  |  |
| Smoking History | 94 (36.8) | 172 (34.3) |
| Never Smoker | 66 (25.9) | 107 (21.4) |
| Former Smoker | 8 (3.1) | 19 (3.8) |
| Current Smoker | 87 (34.1) | 203 (40.5) |
| Unknown |  |  |
| Current Education | 3 (1.2) | 0 |
| No education | 3 (1.2) | 8 (1.6) |
| Eighth grade or less | 6 (2.4) | 7 (1.4) |
| Some high school | 60 (23.5) | 112 (22.4) |
| High school graduate or GED | 23 (9) | 47 (9.4) |
| Vocational or some college | 86 (33.7) | 178 (35.6) |
| College graduate | 38 (14.9) | 55 (11) |
| Graduate school | 36 (14.1) | 94 (18.8) |
| Unknown |  |  |
| Current Employment | 31 (12.2) | 43 (8.6) |
| Unemployed | 2 (0.8) | 5 (1) |
| Student | 95 (37.3) | 188 (37.5) |
| Employed | 3 (1.2) | 7 (1.4) |
| Homemaker | 88 (34.5) | 188 (37.5) |
| Retired | 25 (9.8) | 37 (7.4) |
| Disability | 11 (4.3) | 33 (6.6) |
| Unknown |  |  |
| Current Insurance | 8 (3.1) | 24 (4.8) |
| Free or self pay | 138 (54.1) | 260 (51.9) |
| Commercial | 97 (38) | 194 (38.7) |
| Medicare | 12 (4.7) | 23 (4.6) |
| Medicaid or other government |  |  |
| 2018 Median Family Income in dollars; Median (IQR) | \$103050<br>(\$74432<br>-\$131779) | \$106730<br>(\$83617-<br>\$130450) |
| **Characteristic** | **SSc-ILD**<br>**(n= 255)** | **SSc without ILD**<br>**(n= 501)** |
| **Clinical Features and Care Utilization** |  |  |
| ANA Not Tested | 90 (35.3) | 220 (43.9) |
| ANA Negative | 6 (2.4) | 21 (4.2) |
| ANA Positive | 159 (62.4) | 260 (51.9) |
| Scl-70 Not Tested | 92 (36.1) | 245 (48.9) |
| Scl-70 Negative | 127 (49.8) | 214 (42.7) |
| Scl-70 Positive | 36 (14.1) | 42 (8.4) |
| Charlson Comorbidity Index Including Age, Before Index SSc Code; Median (IQR) | 2 (1-3) | 2 (1-3) |
| Organ Involvement ICD-10 Code Present in Chart | 48 (18.8) | 37 (7.4) |
| AKI | 43 (16.9) | 48 (9.6) |
| CKD | 86 (33.7) | 118 (23.6) |
| Esophageal disease | 160 (62.7) | 203 (40.5) |
| GERD | 62 (24.3) | 48 (9.6) |
| Pulmonary Hypertension |  |  |
| Medication Order in Chart During Study Period | 84 (32.9) | 108 (21.6) |
| Prednisone | 123 (48.2) | 92 (18.4) |
| Mycophenolate |  |  |
| At Least One ED Visit | 110 (43.1) | 169 (33.7) |
| At Least One Hospitalization | 115 (45.1) | 141 (28.1) |

*Definition of abbreviations:* ICD-10 = International Classification of Diseases, Tenth Revision; SSc= systemic sclerosis; ILD= interstitial lung disease; SD = standard deviation; ANA= antinuclear antibody; IQR = interquartile range; AKI= acute kidney injury; CKD= chronic kidney disease; GERD= gastro-esophageal reflux disease; ED= emergency department. Data are n (%) unless otherwise stated.

### ILD Prevalence, and Race/Ethnicity

Among the 756 patients, 501 (66.3%) did not have ILD and 255 (33.7%) had ILD before or during the study period (Table 1). Although there were many differences in organ involvement, laboratory values, and other clinical characteristics between SSc patients with and without ILD, race/ethnicity was the primary demographic characteristic that differed between groups (Table 1). Sensitivity analysis using a more restrictive case definition of ILD, demonstrated similar findings (Table E2). Unadjusted logistic regression models show Black/African American and Asian patients have increased odds of ILD (odds ratio [OR] 1.99, 95% confidence interval [95% CI 1.08, 3.64], p=0.027 and OR 2.53 [95% CI 1.29, 4.93], p=0.007, respectively). After adjusting for SDOH variables, there was a statistically significant association for Asian patients (OR 2.59, [95% CI 1.29, 5.18], p=0.007), (Table E3).

Additionally, there was an increased odds of ILD for both Black/African American and Asian patients in unadjusted sensitivity analyses, which remained significant after adjusting for covariates (Table E3). The effects of other covariates associated with ILD risk are included in Table E4. Further investigation of the functional, clinical, and serological features of Black/African American and Asian patients with ILD is presented in Table E5. On average, from the first available pulmonary function test (PFT), Black/African American patients had restrictive defects (mean percent-predicted FVC at baseline 57% ± 17), with moderate diffusion limitation (mean percent-predicted DLCO at baseline 51% ± 14). Black/African American (n=8, 42%; vs n=5, 25%) and Asian (n=5, 45%; vs n=3, 27%) patients had more anti-RNP positivity than Scl-70.

### Time to ILD Diagnosis

Comparable to differences in having ILD, the timing of an SSc diagnosis, and an ILD diagnosis differed by race/ethnicity; specifically, Black/African American patients were, on average, more likely to have an ILD diagnosis that preceded their SSc diagnosis (59.1%), in contrast to White patients with SSc of whom over 80% were diagnosed with ILD after SSc (p=0.001 for the difference between groups, TableE 6). Similarly, there were differences between racial/ethnic groups in the distribution of time between SSc and ILD diagnoses as shown in Figure 1 (p<0.001, for the difference between groups). For example, Black/African American patients had an ILD code a median of 2.5 months prior to SSc code, in contrast to all other groups of patients. Additionally, Black/African American patients had the shortest maximum time to ILD after SSc, at 3 months. White patients had the widest range of the SSc and ILD interval (maximum of 61 months), while Asian and Hispanic patients had similar maximum time at 44 and 56 months, respectively.

**Figure 1.**
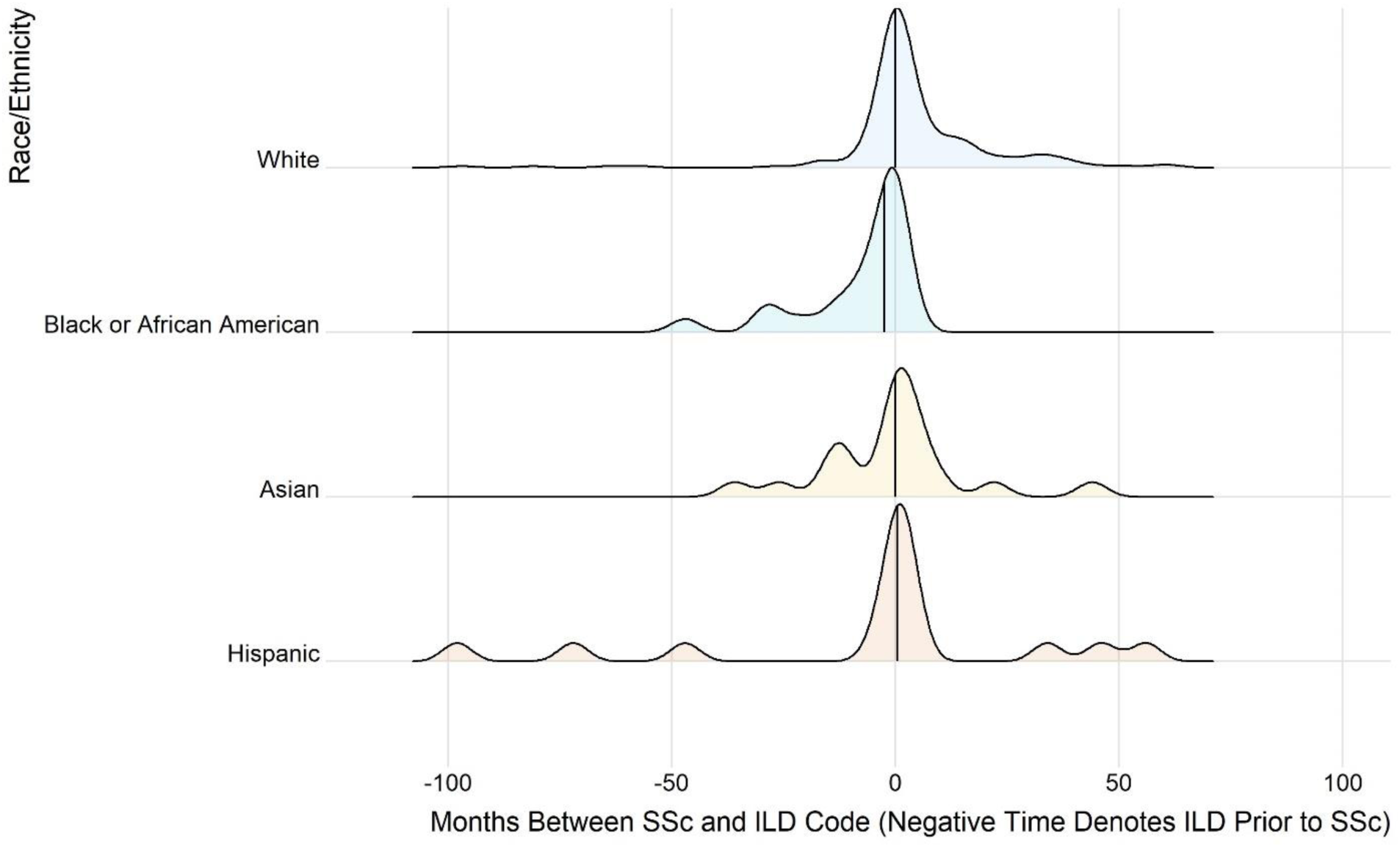
Ridgeline plot of time interval (in months) between systemic sclerosis and interstitial lung disease diagnosis codes by race/ethnicity (unimputed, unknown race/ethnicity not shown). Negative time denotes ILD diagnosis prior to SSc diagnosis. Median in months shown by line for each race/ethnicity. Density plot illustrating distribution of time, not indicating raw data points except for median line. Range and median for each race/ethnicity: White (−166, 61, med = 0mo), Black/African American (−47, 3, med = −2.5mo), Asian (−36, 44, med= 0mo), Hispanic (−98, 56, med=0.5mo). SSc= systemic sclerosis. ILD = interstitial lung disease. Kruskal-Wallis p-value < 0.001 for differences in distribution.

### Age at SSc and SSc-ILD

In addition to the racial/ethnic differences in the timing interval between SSc and ILD, there were significant differences in the age of both SSc and SSc-ILD between racial/ethnic groups. For the entire cohort, the observed average age at ILD diagnosis was 58.5 years; for White patients the average age was 61.5 years and for all non-White patient groups ranged from 46-52.9 years. In the multivariable regression analysis adjusting for SDOH, Black/African American and Hispanic patients were significantly younger for both SSc ([-]9.3 and [-]4.6 years, respectively) and SSc-ILD ([-]10 and [-]6.5 years, respectively) compared to White patients (Table 2). In contrast, the SSc and SSc-ILD age difference between Asian and White patients was substantially attenuated after adjustment (Table 2). The statistically significant younger age at SSc-ILD compared to White patients persisted for Black/African American patients in the sensitivity analysis using the restricted condition (Table E7).

**Table 2.**
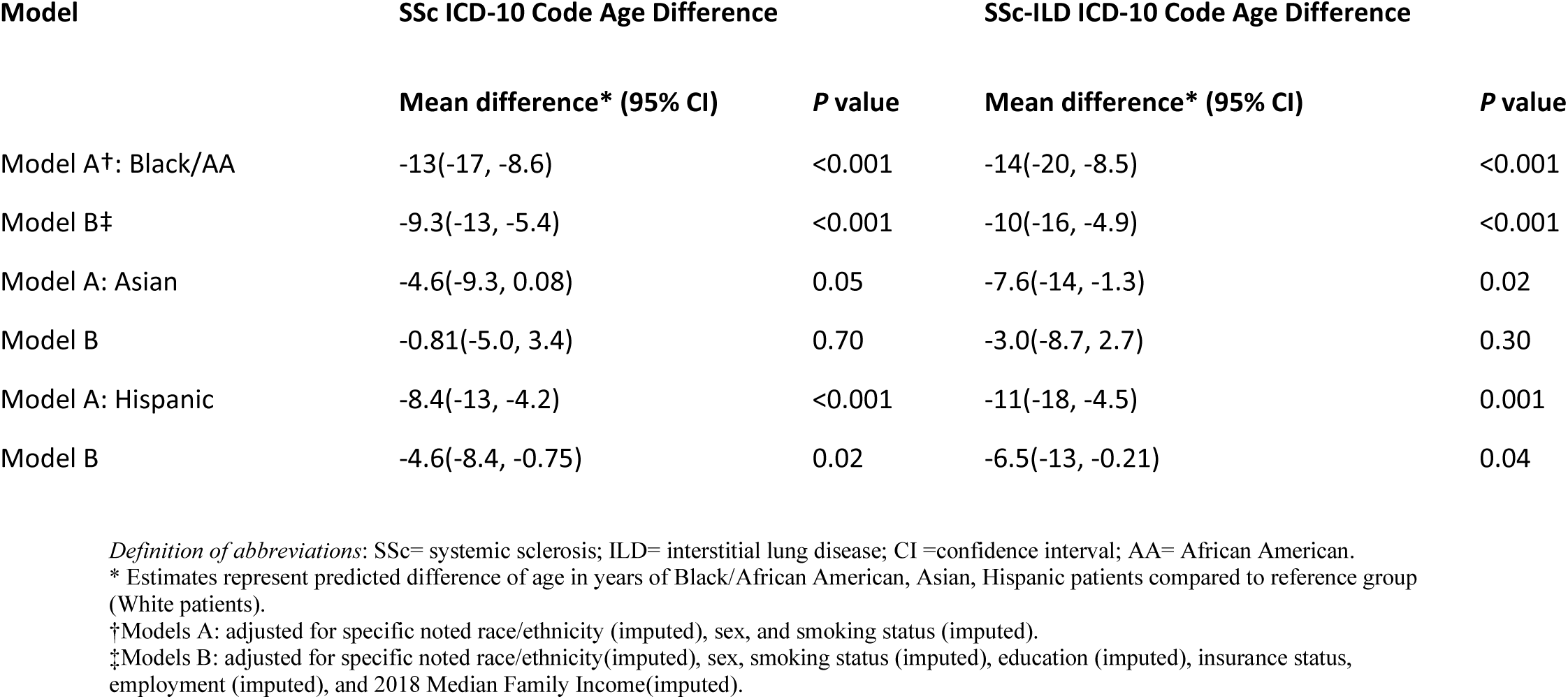
Adjusted mean age difference for age at systemic sclerosis ICD-10 code and systemic sclerosis-interstitial lung disease ICD-10 code, among Black/African American, Asian or Hispanic patients compared to White patients.

| Model | SSc ICD-10 Code Age Difference |  | SSc-ILD ICD-10 Code Age Difference |  |
| --- | --- | --- | --- | --- |
|  | Mean difference* (95% CI) | P value | Mean difference* (95% CI) | P value |
| Model A†: Black/AA | -13(-17, -8.6) | <0.001 | -14(-20, -8.5) | <0.001 |
| Model B‡ | -9.3(-13, -5.4) | <0.001 | -10(-16, -4.9) | <0.001 |
| Model A: Asian | -4.6(-9.3, 0.08) | 0.05 | -7.6(-14, -1.3) | 0.02 |
| Model B | -0.81(-5.0, 3.4) | 0.70 | -3.0(-8.7, 2.7) | 0.30 |
| Model A: Hispanic | -8.4(-13, -4.2) | <0.001 | -11(-18, -4.5) | 0.001 |
| Model B | -4.6(-8.4, -0.75) | 0.02 | -6.5(-13, -0.21) | 0.04 |

*Definition of abbreviations:* SSc= systemic sclerosis; ILD= interstitial lung disease; CI =confidence interval; AA= African American.
\* Estimates represent predicted difference of age in years of Black/African American, Asian, Hispanic patients compared to reference group (White patients).
†Models A: adjusted for specific noted race/ethnicity (imputed), sex, and smoking status (imputed).
‡Models B: adjusted for specific noted race/ethnicity(imputed), sex, smoking status (imputed), education (imputed), insurance status, employment (imputed), and 2018 Median Family Income(imputed).

### Emergency Department Visit and Hospitalization

The mean follow-up time in years by racial/ethnic group was 2.8, 2.6, 3.0, 3.0 for White, Black/African American, Asian, and Hispanic patients respectively. During the study period, 279 (36.9%) patients had at least 1 ED visit, with 990 total visits during the study period. There were 256 patients (33.9%) with at least 1 hospitalization, and 145 (19.2%) had multiple hospitalizations (with 725 total hospitalizations during the study period). By racial/ethnic group, the observed mean number of ED visits and hospitalizations over 3 years average follow-up is shown in Table E8.

Black/African American, and Hispanic patients had over 2-fold increased rate (and approximately 2 for Asian patients) of an ED visit, compared to White patients, even after adjustment for SDOH and clinical variables (Table 3). Notably, patients with ILD had a 60% increased rate of a hospitalization compared to those without ILD (Table E4). Sensitivity analyses examined the possible impact of COVID-19 on hospitalizations, comparing pre and during-COVID-19 periods (Table E9), demonstrating increased rates among non-White patients during the COVID period.

**Table 3.**
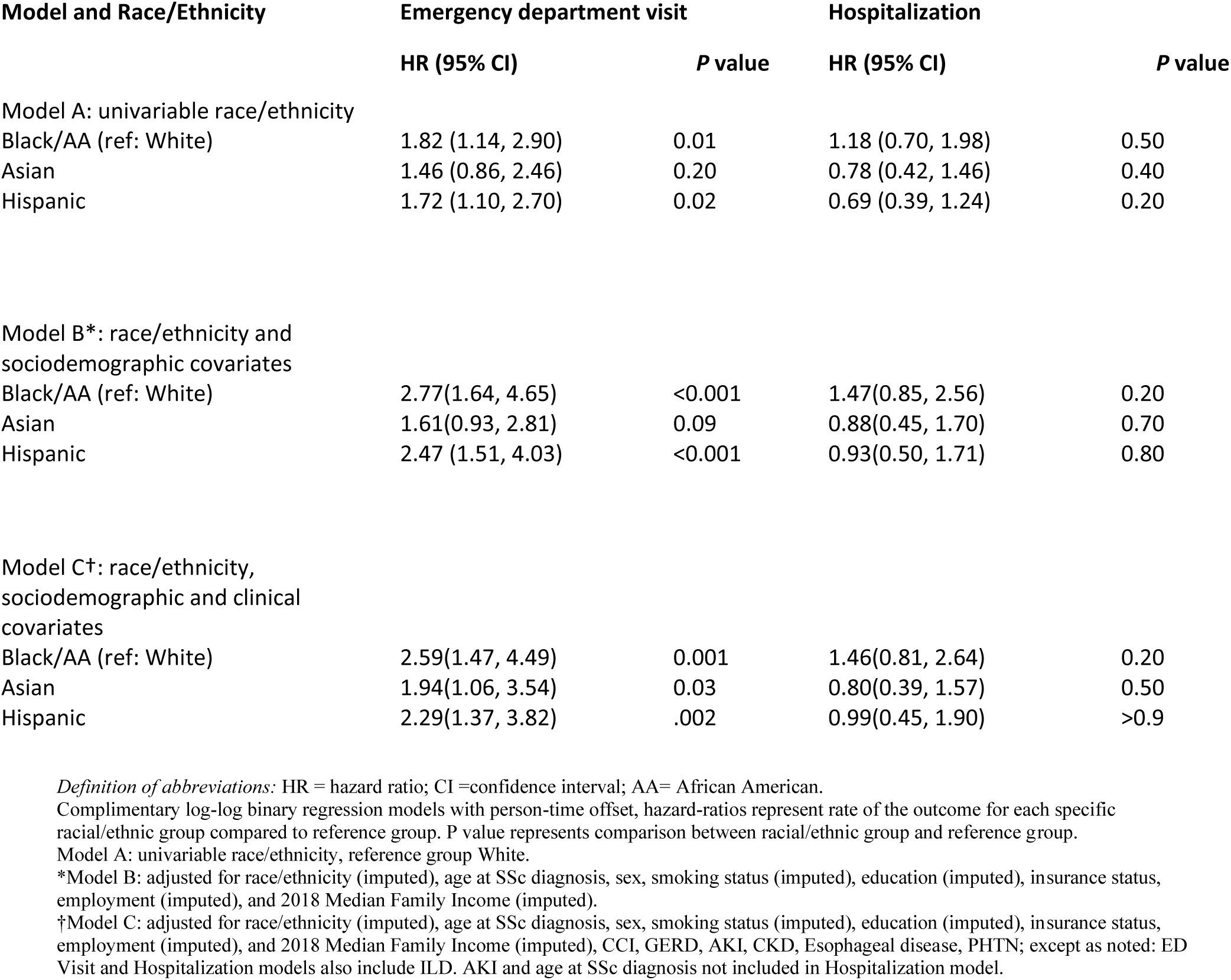
Regression models of race/ethnicity associations with healthcare utilization outcomes: any emergency department visit, any all-cause hospitalization in imputed data.

## Discussion

We found important race/ethnicity differences in the presentation and risk of ILD, its timing with respect to an SSc diagnosis, and important clinical outcomes in a relatively large SSc cohort from a multi-hospital health system. More specifically, in contrast to White patients with SSc, Black/African American patients with SSc were more likely to have ILD, to present at a younger age, to have ILD that precedes a diagnosis of SSc, and to have increased rate of an ED visit. Some, but not all these differences, were noted when comparing Asian and Hispanic patients to White patients with SSc. Additionally, given that Black/African American patients frequently present with ILD preceding a SSc diagnosis, these findings have important implications for general ILD evaluation. These findings suggest that race/ethnicity are important factors to consider in the risk of ILD, and its severity among patients with SSc; extending previously published work by demonstrating that timing of evaluation for minoritized patients is critical.

Other SSc cohorts have had similar demographic findings,^25,26^ and we also found established associations such as GERD with SSc-ILD,^27^ and ILD or PHTN with a hospitalization^28–30^ were demonstrated in this cohort. Although our findings, and those of others, demonstrate that race/ethnicity are important factors associated with presentation and severity of ILD among SSc patients,^8,10,31^ the reasons for this are not clear. We investigated some of the racial/ethnic variability in presentation and outcomes and present novel findings to better understand these differences.

Race is an evolving and poorly defined social construct^32,33^ that impacts health through various mechanisms. Thus, what is captured as differential population distributions of SSc and SSc-ILD is likely a combination of diverse SDOH^34^ and other factors that may correlate with race. Compared to other racial/ethnic groups, Black/African American patients were younger at ILD diagnosis,^11,35,36^ however, there is paucity of data on multi racial/ethnic group comparisons of age at SSc-ILD. We found that compared to White patients, Hispanic and Black/African American patients had significantly younger age at SSc and SSc-ILD diagnosis of 4 to 9 years and 6 to 10 years respectively. Additionally, compared to other groups, more Black/African American patients had ILD prior to SSc, with a very narrow interval between the two diagnoses. The attenuation of age differences between groups by available SDOH data suggests they could explain some of the SSc-ILD age heterogeneity but there may be other relevant exposures or unmeasured factors (eg environmental or genetic) that may explain differences in susceptibility, severity, or phenotypes. Multi-racial/ethnic group differences in adjusted SSc-ILD age have not previously been reported, and may be informative in risk-stratification. The short interval (3 months) between an SSc and an ILD diagnosis for Black/African American patients, coupled with the risk of SSc-ILD earlier in SSc course^37^ supports consideration of earlier pulmonary evaluation; additionally, a higher level of suspicion for SSc in young non-White patients with ILD could be beneficial. These considerations have management and therapeutic implications.^16^

Given significant lung function decline is shown to occur insidiously and earlier in SSc course,^38^ risk stratification and establishing baseline disease severity is important to preserve lung function before irreversible lung damage occurs.^39^ Among the Black/African American patients with available baseline FVC measurement, most were nonsmokers and had moderately severe RLD, which may suggest a more severe phenotype at presentation, aligning with data from prior studies. The baseline FVC and DLCO percentage predicted are similar to baseline values in other cohorts,^40,41^ and non-White patients have been shown to have increased odds of RLD at baseline.^42^ Black and Asian patients have been shown to have lower FVC compared to White patients,^31^ and in one study, African Americans had more extensive ILD at baseline and faster rate of FVC decline compared to other participants.^11^ Our lung function findings, along with other studies suggest clinical importance of obtaining physiological data earlier, and possibly more frequently.

Autoantibodies and environmental exposures^43,44^ could partially explain the variation of SSc-ILD presentation and trajectory. Among African American and Hispanic patients with similar sociodemographic parameters in previous studies, Hispanic patients had higher DLCO, less fibrosis and lower frequency of PAH.^12^ Black/African American patients have been shown to have higher levels of both anti-ribonucleoprotein (U1-RNP) and antifibrillin antibodies (U3-RNP) compared to non-Black patients with SSc,^45–47^ a higher proportion of Asian patients compared to White patients have anti-U1-RNP positivity,^10^ and more severe fibrosis is associated with anti–U1 RNP–positivity.^9^ Among African American patients with scleroderma in the Genome Research in African American Scleroderma Patients (GRASP) cohort, 14-18% were positive for anti-RNPs and 20% had a concomitant overlapping autoimmune disease.^41^ For the subset of Black/African American and Asian patients with ILD in this cohort with testing, over 40% had anti-RNP positivity, most of whom also had additional autoimmune disease considerations. The pathophysiology of autoantibodies and compounded risk from overlapping diseases remains incompletely understood. Regarding possible environmental contributions, in another SSc cohort, overlaying environmental toxin maps in Massachusetts showed hazardous waste and oil disposal sites were significantly associated with higher SSc prevalence, with clustering around neighborhoods with low-income and minority inhabitants.^48^ Additionally, observational studies suggest an association between annual ozone concentration and extensive SSc-ILD imaging severity at diagnosis.^49^ Taken together, it is possible that expanding serological testing, evaluation of environmental factors, and consideration of SSc/SSc-ILD in specific populations, could inform risk assessment and management.

In our study, adjusted analyses show racial/ethnic differences exist in prevalence of ILD and rate of ED visit but not consistently with hospitalization. The limited data on SSc racial/ethnic differences in ED visits and hospitalizations demonstrate increased rates among Black/African American patients compared to White,^18–20^ and pulmonary fibrosis as the condition associated with the highest chance of in-hospital mortality.^20^ Our multi-racial/ethnic analyses expand on differences in ED visits, demonstrating increased significant rates for Black/African American, Hispanic, and Asian patients. Several explanations for increased rates for subpopulations are possible: differences in extent of SSc manifestations independent of adjusted conditions (such as SSc-ILD), contributing comorbidities not fully captured by Charlson Comorbidity Index (CCI), or healthcare delivery (differences in ambulatory/subspeciality access or care coordination throughout health-systems). There was a trend for increased overall and COVID-19 period hospitalization rates for Black/African American patients (which has been noted in PF in general^36^); overall, differences in hospitalization may not have been observed due to sample size, low acuity of ED visit leading to hospitalization, random effects such as care transition (ie, transfer to another hospital for admission) or other features not captured.

Inaccessible or dissimilar care has been described as a potential explanation for racial/ethnic differences in SSc overall. Longitudinal cohort studies^13^ and clinical trials,^11^ where comparable access and management is more likely, suggest that some endpoint disparities may be minimized under these conditions. However, race/ethnicity and socio-economic status data have not been reported in most SSc randomized trial publications,^50^ and there are other unmeasured contributing factors that can aid in comprehensively studying disparities. Reducing disparities has important clinical implications, fully examining the burden of SSc-ILD in clinical practice provides expanded care opportunities, which future EHR studies can facilitate.

This study has several limitations. One common limitation in secondary data analyses, the true duration of disease (or date from first non-Raynaud symptom) cannot be measured with certainty, which is why we used first-time use of the ICD-10 code as a proxy for “index” disease date. Code misclassification is another known limitation in EHR and claims data,^51^ however, similar findings were demonstrated in the restricted sensitivity analyses based on a clinician adding ILD to the problem list, albeit there are overall limitations of ILD coding.^17,52^ Conversely, patients that may have ILD may not have been captured, either due to lack of evaluation or coding for ILD. Disease activity, characteristics of SSc (ie, subtype), medication use (timing of initiation and treatment response), cause of ED visit or hospitalization and other information that may further explain some of these findings are unknown, however we included a wide number of available SDOH data in this study. Local and geographical differences between patients is not available, and this is a multihospital single health system study which may not be broadly generalizable, further validation in other health systems is necessary. There may be variability in PFT reference equation calculation, and race/ethnicity is self-reported therefore confirming accuracy in chart documentation is limited. As some were not available in our system, CT images were not independently reviewed.

### Conclusions

In this study, we confirmed known associations with SSc-ILD, and add information on racial/ethnic differences in age and timing of diagnosis of SSc-ILD, particularly that non-White patient groups were younger. More Black/African American patients were diagnosed with SSc-ILD before SSc and at a younger age, and at baseline, had diminished lung function, with more anti-RNP positivity than Scl-70 positivity. Additionally, non-White patients had increased rates of an ED visit. Multiple factors may drive differential disease timing and outcomes in non-White patient groups, and further investigation is warranted. ILD is an early manifestation and leading cause of death in SSc, and earlier identification is critical for improving outcomes.

## Data Availability

Data is not currently publicly available but may be available upon reasonable request to the authors.

## Acknowledgements

The authors thank Drs. John E. Orav and Stuart Lipsitz for their expertise.

This work was conducted with support from Harvard Catalyst | The Harvard Clinical and Translational Science Center (National Center for Advancing Translational Sciences, National Institutes of Health Award UL1 TR002541) and financial contributions from Harvard University and its affiliated academic healthcare centers. The content is solely the responsibility of the authors and does not necessarily represent the official views of Harvard Catalyst, Harvard University and its affiliated academic healthcare centers, or the National Institutes of Health.

## Additional information

The e-Tables are available online under “Supplementary Data.”

