## Supplemental File for "Racial and ethnic associations with interstitial lung disease and healthcare utilization in patients with systemic sclerosis"

ONLINE DATA SUPPLEMENT

| **Table E1:** Selected ICD-10 code used for disease processes | |
|---|---|
| Disease | ICD-10 Code Used |
| Systemic sclerosis | M34* |
| Interstitial lung disease | J84* |
| Gastro-esophageal reflux disease (GERD) | K21.9 |
| Esophageal disease | K22.4 |
| Raynaud | I73* |
| Pulmonary hypertension (PHTN) | I27.20, I27.29 |
| Chronic kidney disease (CKD) | N18* |
| Acute kidney injury (AKI) | N17.9 |

### Results

**Table E2:** Sensitivity analysis comparison of selected baseline characteristics of patients with ILD on problem list vs not on problem list

| Characteristic | ILD Code Problem List N= 133 | ILD Code Not in Problem List N=623 | P value |
|---|---|---|---|
| Age at Index SSc Code, years, mean ± SD | 58.2 (13.9) | 59.6 (14.8) | .308 |
| Female | 100 (75.2) | 532 (85.4) | .006 |
| White | 97 (72.9) | 510 (81.9) | <.001 |
| Black | 16 (12) | 29 (4.6) | |
| Hispanic | 6 (4.5) | 40 (6.4) | |
| Asian | 13 (9.8) | 24 (3.8) | |
| Unknown | 1 (0.75) | 20 (3.2) | |
| Never Smoker | 54 (40.6) | 40 (32.8) | .639 |
| Former Smoker | 32 (24.1) | 34 (27.9) | |
| Current Smoker | 4 (3) | 4 (3.3) | |
| Unknown | 43 (32.3) | 44 (36.1) | |
| **Clinical Features and Care Utilization** | | | |
| Scl-70 Not Tested | 56 (42.1) | 281 (45.1) | .155 |
| Scl-70 Negative | 57 (42.9) | 284 (45.6) | |
| Scl-70 Positive | 20 (15) | 58 (9.3) | |
| Charlson Comorbidity Index Including Age, Before Index SSc Code; Median (IQR) | 2 (1-3) | 2 (1-3) | .194 |
| Organ Involvement Code Present in Chart | | | |
| Esophageal disease | 54 (40.6) | 150 (24.1) | <.001 |
| GERD | 89 (66.9) | 274 (44) | <.001 |
| Pulmonary Hypertension | 37 (27.8) | 73 (11.7) | <.001 |
| Treatment During Study Period | | | |
| Prednisone | 48 (36.1) | 144 (23.1) | .003 |
| Mycophenolate | 78 (58.6) | 137 (21.9) | <.001 |

*Definition of abbreviations*: SSc= systemic sclerosis; ILD= interstitial lung disease; SD = standard deviation; IQR = interquartile range; GERD= gastro-esophageal reflux disease.

Data are n (%) unless otherwise stated.

**Table E3.** Logistic regression models for race/ethnicity and interstitial lung disease

| Model and Race/Ethnicity | ILD ICD-10 Code | | ILD ICD-10 code on problem list* | |
|---|---|---|---|---|
| | OR (95% CI) | P value | OR (95% CI) | P value |
| Model A†: univariate race/ethnicity | | | | |
| Black/AA (ref: White) | 1.99(1.08, 3.64) | .027 | 2.85 (1.50, 5.45) | 0.002 |
| Asian | 2.53 (1.29, 4.93) | .007 | 2.92 (1.44, 5.92) | 0.003 |
| Hispanic | 1.13 (0.60, 2.13) | 0.70 | 0.79 (0.32, 1.90) | 0.60 |
| Model B‡: race/ethnicity and sociodemographic covariates | | | | |
| Black/AA (ref: White) | 1.78(0.93, 3.41) | 0.084 | 2.73 (1.34, 5.57) | 0.006 |
| Asian | 2.59 (1.29, 5.18) | 0.007 | 2.76 (1.31, 5.80) | 0.008 |
| Hispanic | 1.10 (0.56, 2.15) | 0.80 | 0.81 (0.32, 2.04) | 0.70 |

Definition of abbreviations: ICD-10 = International Classification of Diseases, Tenth Revision; ILD= interstitial lung disease; OR = odds ratio; AA = African American.
*Sensitivity analysis restricting to those with interstitial lung disease code on problem list.
Model A: univariate analysis of race/ethnicity and ILD, reference group White.
†Model B: adjusted for race/ethnicity(imputed), age at SSc diagnosis, sex, smoking status (imputed), education (imputed), insurance status, employment (imputed) and 2018 Median Family Income (imputed).

**Table E4.** Multivariable models of interstitial lung disease diagnosis, any emergency department visit, any all-cause hospitalization in imputed data.

| Variable | ILD* | | ED Visit† | | Hospitalization† | |
|---|---|---|---|---|---|---|
| | OR (95% CI) | *P* value | HR (95% CI) | *P* value | HR (95% CI) | *P* value |
| Age at index SSc code, years | 1.00 (0.98, 1.02) | 0.8 | 0.98 (0.97, 1.00) | 0.011 | | |
| Male sex (ref female) | 1.44 (0.94, 2.22) | 0.093 | 0.82 (0.56, 1.20) | 0.3 | 1.13 (0.79, 1.63) | 0.5 |
| Black/African American (ref White) | 2.00 (1.02, 3.95) | 0.045 | 2.59 (1.46, 4.59) | 0.001 | 1.46 (0.81, 2.64) | 0.2 |
| Asian | 2.29 (1.10, 4.77) | 0.027 | 1.93 (1.06, 3.53) | 0.032 | 0.80 (0.41, 1.57) | 0.5 |
| Hispanic | 1.09 (0.55, 2.20) | 0.8 | 2.29 (1.37, 3.84) | 0.002 | 0.99 (0.52, 1.90) | >0.9 |
| Former Smoker (ref never smoker) | 1.33 (0.88, 2.01) | 0.2 | 1.46 (1.05, 2.02) | 0.024 | 0.99 (0.68, 1.44) | >0.9 |
| Current Smoker | 1.01(0.46, 2.25) | >0.9 | 1.57 (0.82, 2.98) | 0.2 | 1.11 (0.57, 2.17) | 0.8 |
| College degree or higher | 1.09(0.76, 1.58) | 0.6 | 0.97 (0.73, 1.30) | 0.9 | 0.71 (0.51, 0.97) | 0.031 |
| Employed/Retired | 0.83(0.48, 1.43) | 0.5 | .050 (0.32, 0.77) | .002 | 0.49 (0.31, 0.76) | 0.002 |
| Self-pay (ref Commercial) | 0.73(0.30, 1.76) | 0.5 | 0.95 (0.46, 1.97) | 0.9 | 0.70 (0.31, 1.56) | 0.4 |
| Medicare | 1.10(0.74, 1.63) | 0.6 | 0.97 (0.70, 1.34) | 0.8 | 1.02 (0.74, 1.39) | >0.9 |
| Medicaid or other government | 0.77(0.34, 1.76) | 0.5 | 1.01 (0.52, 1.98) | >0.9 | 1.14 (0.54, 2.39) | 0.7 |
| 2018 Median Income, dollars | 1.00(1.00, 1.00) | 0.3 | 1.00 (1,00, 1.00) | 0.015 | 1.00 (1,00, 1.00) | 0.15 |
| Charlson Comorbidity Index | 0.90(0.78, 1.05) | 0.2 | 1.35 (1.22, 1.50) | <.001 | 1.40 (1.28, 1.53) | <.001 |
| GERD | 2.60(1.82, 3.72) | <.001 | 2.30 (1.69, 3.21) | <.001 | 1.95 (1.42, 2.67) | <.001 |
| AKI | | | 6.48 (3.94, 10.7) | <.001 | | |
| CKD | | | .70 (0.43, 1.13) | 0.14 | 1.79 (1.24, 2.57) | 0.002 |

| | | | | | | |
|---|---|---|---|---|---|---|
| Esophageal disease | 1.12(0.76, 1.65) | 0.60 | 0.88 (0.64,1.21) | 0.4 | 0.69 (0.49, 0.96) | 0.027 |
| SCl-70 Positive (ref Nontested) | 1.63(1.16, 2.30) | .005 | | | | |
| Scl-70 Negative (ref Nontested) | 2.49(1.45, 4.27) | <.001 | | | | |
| Pulmonary Hypertension | | | 0.75 (0.50, 1.14) | 0.2 | 1.77 (1.24, 2.52) | 0.002 |
| ILD | | | 0.88 (0.65, 1.19) | 0.4 | 1.60 (1.19, 2.15) | 0.002 |

*Definition of abbreviations*: OR = odds ratio; HR = hazard ratio; SSc= systemic sclerosis; ILD= interstitial lung disease; SD = standard deviation; IQR = interquartile range; AKI= acute kidney injury; CKD= chronic kidney disease; GERD= gastro-esophageal reflux disease; ED= emergency department.

*Logistic regression model

† Complimentary log-log model

Each outcome model adjusted for covariates with values in the rows: race/ethnicity(imputed), age at SSc diagnosis, sex, smoking status (imputed), education (imputed), insurance status, employment (imputed), and 2018 Median Family Income(imputed), CCI, GERD, CKD, Esophageal disease, Scl70 testing. Rows without data do not include that covariate in model. Hospitalization model reflects overall HR during entire study period (pre and during COVID periods)

**Table E5A.** Functional, serological and clinical characteristics of Black/African American patients with systemic sclerosis interstitial lung disease

| | Baseline FVC† Mean ±SD | Baseline DLCO‡ Mean ±SD | GERD | Anti-topoisomerase I Positive Among Tested, N=20 | Anti-RNP Positive Among Tested, N=19 | Pulmonary Hypertension Noted | Mycophenolate Use Ever Noted | End Stage Lung Disease | Any Hospitalization |
|---|---|---|---|---|---|---|---|---|---|
| N*(%), Mean | 57% Predicted ± 17 | 51% Predicted ± 14 | 17 (94%) | 5 (25%) | 8 (42%) | 7 (38%) | 16 (88%) | 2 status post bilateral transplant | 10 (55%) |

*Definition of abbreviations*: FVC=forced vital capacity; DLCO= diffusing capacity for carbon monoxide; GERD= gastro-esophageal reflux disease; RNP=antinuclear ribonucleoprotein. RNP unspecified, n=1 U1-RNP.
N=24 with ILD code; N=18 with code validation during study with additional 1 after study period
(PPV 79% during; 83% for PPV after study period).
*N and % are among n=18 with code validation unless otherwise noted.
N=12 with available PFT measurement noted. N=11 nonsmokers per chart.
†: n=11 with available baseline FVC – first available in system per note or report.
‡: n=7 with available baseline DLCO – first available in system per note or report.
7 of 8 patients with anti-RNP positivity had notation with concern for MCTD, SLE, inflammatory myopathy with SSc overlap.

**Table E5B.** Functional, serological and clinical characteristics of Asian patients with systemic sclerosis interstitial lung disease

| | Percent-predicted FVC at baseline † Mean ±SD | Percent-predicted DLCO at baseline ‡ Mean ±SD | GERD | Anti-topoisomerase I Positive Among Tested, N=11 | Anti-RNP Positive Among Tested, N=11 | Pulmonary Hypertension Noted | Mycophenolate Use Ever Noted | End Stage Lung Disease | Any Hospitalization |
|---|---|---|---|---|---|---|---|---|---|
| N*(%), Avg | 69% Predicted ± 21 | 55% Predicted ± 21 | 13 (87%) | 3 (27%) | 5 (45%) | 5 (33%) | 11 (73%) | 0 | 5 (33%) |

*Definition of abbreviations*: Avg: average. FVC=forced vital capacity. DLCO= diffusion capacity of carbon monoxide. GERD= gastro-esophageal reflux disease. RNP=antinuclear ribonucleoprotein.
N=20 with ILD code; N=15 with code validation during study (PPV 75% during study period).
*N and % are among n=15 with code validation unless otherwise noted.
N=15 with available PFT measurement noted. N=12 nonsmokers per chart.
†: n=15 with available baseline FVC and FEV1/FVC >70 – first available in system per note or report
‡: n=14 with available baseline DLCO – first available in system per note or report
RNP unspecified, n=1 U1-RNP.
4 of 5 patients had concern for MCTD, Sjogren’s, inflammatory myopathy with SSc overlap.

**Table E6.** Racial/ethnic distribution of patients by timing of interstitial lung disease ICD-10 code compared to timing of systemic sclerosis ICD-10 code

| Race/ Ethnicity | Total with ILD (Before, During Study Period, n) | ILD Before SSc (n= 64) | ILD Same Date or After SSc (n=188) |
|---|---|---|---|
| Asian | 20 | 7 (35) | 13 (65) |
| Black/AA | 22 | 13 (59) | 9 (41) |
| Hispanic | 16 | 4 (25) | 12 (75) |
| White | 194 | 40 (21) | 154 (80) |

*Definition of abbreviations*: ILD = interstitial lung disease; SSc = systemic sclerosis; AA = African American.
Data are n (%) unless otherwise stated.
Non-imputed observed data, unknown race/ethnicity not shown, n=3. Percent is per row. Comparison based on Fisher exact test, p-value = 0.0013.

**Table E7.** Age difference for systemic sclerosis-interstitial lung disease for Black/African American, Asian or Hispanic patients compared to White patients, among patients with ILD on problem list

| Model | SSc-ILD Age Difference | |
|---|---|---|
| | Estimates* (95% CI) | *P* value |
| Model A†: Black/AA | -14(-23, -6.1) | <0.001 |
| Model B‡ | -10(-18, -2.5) | 0.010 |
| Model A: Asian | -5.0(-13, 3.4) | 0.2 |
| Model B | -0.76(-8.2, 6.6) | 0.8 |
| Model A: Hispanic | -13(-25, -1.6) | 0.026 |
| Model B | -4.4(-15, 6.0) | 0.4 |

*Definition of abbreviations*: ICD-10 = International Classification of Diseases, Tenth Revision; SSc= systemic sclerosis; ILD= interstitial lung; AA = African American.

*Sensitivity analysis - estimates represent predicted difference of age in years of Black/African American, Asian, Hispanic patients separately compared to reference group (White patients) among patients with ILD on problem list.

†Models A: adjusted for specific race or ethnicity(imputed), with sex and smoking status (imputed).

‡Models B: adjusted for specific race or ethnicity(imputed), with sex and smoking status (imputed), education (imputed), insurance status, employment (imputed) and 2018 Median Family Income(imputed).

**Table E8.** Mean ED visit and hospitalizations by racial/ethnic groups over 3 year average followup, in unimputed data

| Race/ Ethnicity | Mean number of ED visits | Mean number of hospitalizations |
|---|---|---|
| Asian | 1.60 | 1 |
| Black/AA | 2.10 | 1.5 |
| Hispanic | 1.60 | 0.8 |
| White | 1.32 | 1.05 |

*Definition of abbreviations*: ED = emergency department; AA = African American.

**Table E9.** Models of race/ethnicity associations with any all-cause hospitalization in imputed data, before (January 2016-February 2020) and during COVID 19 (March 2020-June 2021)

| **Model and Race/Ethnicity** | **Hospitalization Pre COVID*** | | **Hospitalization During COVID†** | |
|---|---|---|---|---|
| | **HR (95% CI)** | ***P* value** | **HR (95% CI)** | ***P* value** |
| Model A: univariable race/ethnicity | | | | |
| Black/AA (ref: White) | 0.94 (0.47, 1.89) | 0.90 | 1.76 (0.83, 3.72) | 0.14 |
| Asian | 0.60 (0.26, 1.38) | 0.20 | 1.16 (0.46, 2.94) | 0.70 |
| Hispanic | 0.42 (0.17, 1.04) | 0.06 | 1.11 (0.47, 2.60) | 0.80 |
| Model B: race/ethnicity and sociodemographic covariates | | | | |
| Black/AA (ref: White) | 1.12 (0.54, 2.31) | 0.80 | 2.32 (1.02, 5.25) | 0.04 |
| Asian | 0.65 (0.27, 1.53) | 0.30 | 1.49 (0.57, 3.91) | 0.40 |
| Hispanic | 0.53 (0.21, 1.33) | 0.20 | 1.51 (0.61, 3.72) | 0.40 |
| Model C: race/ethnicity, sociodemographic and clinical covariates | | | | |
| Black/AA (ref: White) | 1.07 (0.50, 2.26) | 0.90 | 2.39 (1.04, 5.48) | 0.04 |
| Asian | 0.54 (0.22, 1.30) | 0.20 | 1.44 (0.55, 3.76) | 0.50 |
| Hispanic | 0.62 (0.24, 1.62) | 0.30 | 1.67 (0.68, 4.13) | 0.30 |

*Definition of abbreviations*: CI =confidence interval; AA= African American.
Complimentary log-log binary regression models with person-time offset, hazard ratios represent rate of the outcome for each specific racial/ethnic group compared to reference group. P value represents comparison between racial/ethnic group and reference group.
* Models with participants (offset with person time up to March 2020) and hospitalizations before March 2020. HR represents pre-pandemic hospitalization baseline rate comparisons.

† Models with all participants (offset with overall person time), with hospitalizations March 2020-June 2021. HR represents hospitalization during COVID rate comparison.
Model A: univariable race/ethnicity, reference group White.
Model B: adjusted for race/ethnicity (imputed, sex, smoking status (imputed), education (imputed), insurance status, employment (imputed), and 2018 Median Family Income (imputed).
Model C: adjusted for race/ethnicity (imputed), sex, smoking status (imputed), education (imputed), insurance status, employment (imputed), 2018 Median Family Income (imputed), CCI, GERD, CKD, Esophageal disease, ILD, PHTN.
